# Evaluating the Safety and Efficacy of Topical Insulin for Ocular Disease: A Systematic Review

**DOI:** 10.1101/2024.02.24.24303321

**Authors:** Luís Jesuino de Oliveira Andrade, Gabriela Correia Matos de Oliveira, Caroline Santos França, Luís Matos de Oliveira

## Abstract

**Introduction:** Ocular surface disorders are prevalent, impacting millions worldwide and causing significant morbidity. Conventional treatments often fall short in addressing refractory cases. Topical insulin has emerged as a potential therapeutic option.

**Objective:** This systematic review aimed to evaluate the safety and efficacy of topical insulin for ocular diseases.

**Methods:** We conducted a systematic review in major databases including the PubMed, MEDLINE, and EMBASE for studies published from (1976 Jan - 2024 Feb) investigating topical insulin for ocular conditions. Studies were screened and selected based on predefined inclusion/exclusion criteria. Data on safety and efficacy were extracted and analyzed.

**Results:** Ten studies (1 case-control, 3 randomized prospective, 3 retrospective, and 3 double-blind designs) met the inclusion criteria. Studies explored various indications, including neurotrophic corneal ulcers, persistent epithelial defects, recurrent epithelial erosions, dry eye disease, and postoperative corneal wound healing in diabetic patients. Overall, findings suggested promising outcomes with topical insulin: promoting healing of refractory neurotrophic corneal ulcers, accelerating reepithelialization in persistent epithelial defects, reducing recurrence of recurrent epithelial erosions, improving symptoms and reducing corneal staining in dry eye disease, enhancing postoperative corneal epithelial wound healing in diabetic patients. Adverse events were minimal and primarily reported as transient stinging or discomfort.

**Conclusion:** This review provides encouraging evidence for the therapeutic potential of topical insulin in diverse ocular diseases. While methodological limitations exist, particularly in non-randomized studies, the current body of literature suggests topical insulin may offer a safe and effective treatment option for patients with refractory corneal disorders.

## INTRODUCTION

Ocular conditions encompass a wide spectrum of diseases, which pose significant challenges to patients and clinicians’ alike.^1^ These conditions often lead to vision impairment and blindness, necessitating the exploration of novel therapeutic approaches to mitigate their impact on individuals’ quality of life. The eye is a complex organ with a unique physiology, and the treatment of ocular conditions often requires targeted, localized therapy. Topical administration, in the form of eye drops, is a common method of drug delivery for ocular conditions.^2^

Insulin, a well-established hormone primarily known for its role in glucose metabolism, has garnered attention for its potential pleiotropic effects in ocular tissues. Beyond its traditional role in diabetes management, insulin has been recognized for its anti-inflammatory, anti-oxidative, and wound healing properties within the context of ocular pathologies.^3^ The multifaceted functions of insulin present an intriguing opportunity to investigate its therapeutic potential in the management of ocular conditions, potentially offering a new avenue for treatment.

The topical administration of insulin has emerged as a promising modality for delivering the hormone directly to ocular tissues, bypassing systemic adverse effects associated with traditional routes of administration.^4^ By targeting the affected ocular tissues locally, topical insulin may exhibit enhanced efficacy while minimizing systemic exposure, minimizing side effects, and providing a targeted delivery to the affected tissues.^5,6^

Despite the potential benefits of topical insulin for ocular diseases, its clinical use remains limited. There is a significant gap in knowledge regarding the safety and efficacy of this approach, particularly in terms of long-term outcomes and optimal treatment protocols.^4,7^ Thus, systematic reviews and meta-analyses of existing evidence hold the key to addressing the knowledge gap surrounding the use of ocular topical insulin. By comprehensively assessing its safety and efficacy in treating various ocular diseases, these rigorous assessments can inform clinical practice and guide future research directions.

This systematic review aims to provide a comprehensive evaluation of the safety and efficacy of topical insulin for ocular conditions.

## METHOD

### 1. Search strategy

A systematic review was conducted following the Preferred Reporting Items for Systematic Reviews and Meta-Analyses (PRISMA) guidelines.^8^ The research strategy involved key electronic databases including PubMed, Scopus, Embase, and Web of Science from their 1976 Jan - 2024 Feb. The search included Medical Subject Headings (MeSH) terms and keywords “topical insulin” and “ocular disease”.

### 2. Eligibility criteria

We conduct a systematic review and meta-analysis that evaluated the safety and efficacy of topical insulin for ocular conditions. Studies were eligible for inclusion if they meet the following criteria: randomized controlled trial design, investigated the use of topical insulin for any ocular condition, compared topical insulin to another treatment (placebo, standard of care) or different doses/formulations of topical insulin, reported safety outcomes (adverse events) and/or efficacy outcomes (visual acuity, intraocular pressure). Our data selection process focused solely on comprehensive research articles and established studies accessible in peer-reviewed medical publications. We deliberately excluded information sourced from abridged summaries, retrospective surveys, editorial viewpoints, isolated patient experiences, and individual communications.

### 3. Study selection and data extraction

Data from eligible studies were extracted using a standardized data extraction form. The following data will be extracted:

➢ Human studies investigating the safety and efficacy of topical insulin for ocular disease
➢ Studies published in the English language.
➢ Study characteristics: study design, sample size, population, and methods.
➢ Intervention characteristics: type of topical insulin, dose, frequency of administration.
➢ Comparison intervention: type of comparator, dose, frequency of administration.
➢ Outcome measures: safety outcomes and efficacy outcomes.

### 4. Data Synthesis

After an extensive search and de-duplication, an initial screening of all articles was performed based solely on title and abstract. Subsequently, a comprehensive review of the full texts of the articles included in the initial stage was conducted for final inclusion. We utilized a standardized data spreadsheet to extract outcome measures from text, tables, and figures in human studies. The assessment of bias risk in human studies was carried out using the Quality Assessment Tools provided by the National Institutes of Health (NIH).^9^

In lieu of conducting a meta-analysis, the authors elected to perform a descriptive review of the data. This decision was made in light of the substantial heterogeneity observed across the appraised studies.

### 5. Analyzing findings and reaching a conclusion

The findings from the systematic review were presented in a descriptive style, analyzed, and final conclusions were drawn.

The potential for bias was evaluated for each study included by utilizing the Quality Assessment of Diagnostic Accuracy Studies (QUADAS)-2 risk assessment tool.^10^

## RESULTS

Our systematic search for relevant studies employed a PRISMA flow diagram (Figure 1) to transparently illustrate the selection process. This meticulous approach commenced with the identification of 146 potential articles through targeted electronic database searches utilizing carefully crafted search terms. Each identified article was then rigorously assessed by title and abstract for relevance. Following deduplication, 47 articles progressed to the next stage, involving a meticulous screening of both abstracts and full texts. This in-depth evaluation ensured that only studies explicitly investigating the safety and efficacy of topical insulin for ocular disease were included. Ultimately, 10 articles^(11-20)^ met the predefined inclusion criteria and were deemed worthy of further analysis and data extraction.

**Fig. 1.**
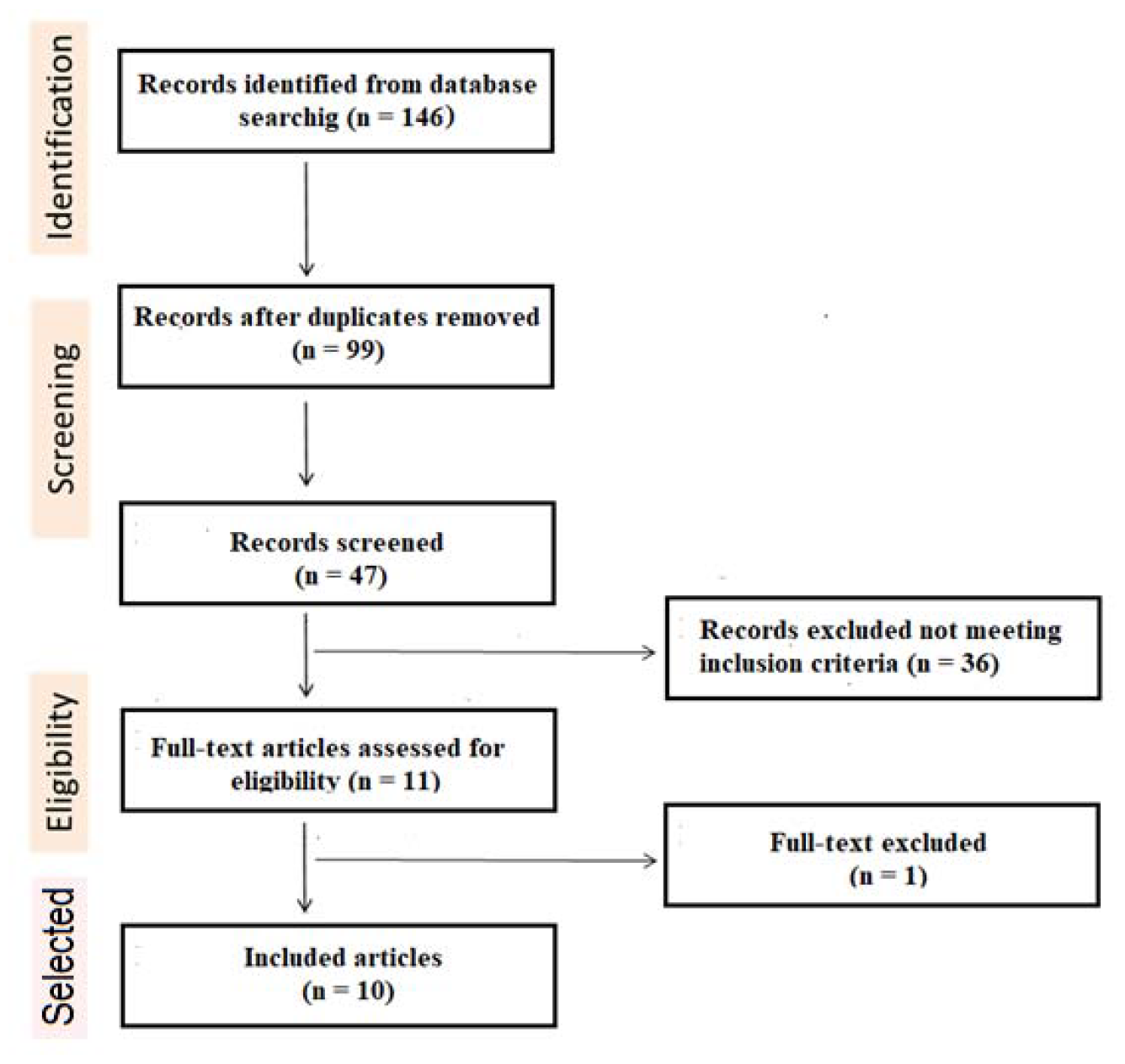
Flow chart on the selection process of eligible studies.

A comprehensive review of the literature explored the safety and efficacy of topical insulin in managing ocular pathologies. Ten studies met the inclusion criteria, encompassing a variety of methodologies such as case-control (one study), randomized prospective (three studies), retrospective (three studies), and double-blind designs (three studies). These investigations were geographically diverse and included sample sizes ranging from 3 to 160 participants.

### Summarizing the main features of the included studies, including their objectives, methods, and conclusion

➢ Reference: Wang AL, et al.^11^; Purpose: “To report the clinical course of 6 patients with refractory neurotrophic corneal ulcers that were treated with topical insulin drops”; Type of study: Retrospective study; Conclusion: “Topical insulin may be a simple and effective treatment for refractory neurotrophic corneal ulcers”.
➢ Reference: Diaz-Valle D, et al.^12^; Purpose: “To investigate the effect of topical insulin on epithelization in persistent epithelial defects refractory to usual treatment compared to autologous serum”; Type of study: Retrospective, consecutive case-control series; Conclusion: “Topical insulin is an effective treatment and safely promotes healing of persistent epithelial defects”.
➢ Reference: Diaz-Valle D, et al.^13^; Purpose: “To evaluate insulin eye drops for persistent epithelial defects that are refractory to usual treatment in clinical practice and to analyze how it may improve epithelization”; Type of study: Prospective non-randomized; Conclusion: “Topical insulin can promote and accelerate corneal reepithelization of refractory persistent epithelial defects”.
➢ Reference: Esmail A, et al.^14^; Purpose: “To evaluate the effect of topical insulin in treatment of recurrent epithelial corneal erosion”; Type of study: Prospective non-randomized; Conclusion: “Topical insulin can promote corneal reepithelization in recurrent epithelial erosion and decreases recurrence in these cases”.
➢ Reference: Burgos-Blasco B, et al.^15^; Purpose: “To evaluate the efficacy of insulin eye drops for dry eye disease in reducing corneal staining and improving symptoms”; Type of study: Retrospective case series; Conclusion: “The excellent results presented in these case series illustrate topical insulin as a promising treatment in dry eye disease with refractory epithelial damage”.
➢ Reference: Soares RJDSM, et al.^16^; Purpose: “To evaluate the clinical outcome of patients with refractory neurotrophic keratopathy in stages 2 and 3 treated with topical insulin”; Type of study: Retrospective study; Conclusion: “Our results suggest that topical insulin drops may be an effective therapeutic in refractory neurotrophic keratopathy”.
➢ Reference: Balal S, et al.^17^; Purpose: “To evaluate the effectiveness of insulin eye drops for treating refractory persistent epithelial defects”; Type of study: Prospective case series; Conclusion: “The use of topical insulin eye drops led to a successful resolution of persistent epithelial defects”.
➢ Reference: Dasrilsyah AM, et al.^18^; Purpose: “To measure and compare the effect of topical insulin (0.5 units, 4 times per day) versus artificial tears (Vismed, sodium hyaluronate 0.18%, 4 times per day) for the healing of postoperative corneal epithelial defects induced during vitreoretinal surgery in diabetic patients”; Type of study: Double-blind randomized controlled; Conclusion: “Topical insulin is more effective compared with artificial tears for the healing of postoperative corneal epithelial defects induced during vitreoretinal surgery in diabetic patients, without any adverse events”.
➢ Reference: Fai S, et al.^19^; Purpose: “To determine the effect of topical insulin of 3 concentrations on postoperative corneal epithelial wound healing in diabetic patients”; Type of study: Double blind randomized controlled; Conclusion: “Topical insulin 0.5 units 0.5, 1, and 2 units per drop 4 times per day is most effective for healing corneal epithelial defect in diabetic patients after vitrectomy surgery compared with placebo and higher concentrations”.
➢ Reference: Aniah Azmi N, Bastion MC.^20^; Purpose: “To assess the short-term effects of topical insulin 1 unit/drop 4 times per day for 4 weeks on the symptoms and signs of diabetic with dry eye disease”; Type of study: Randomized, double-blind interventional study; Conclusion: “The study has shown a significant and similar improvement in the Ocular Surface Disease Index score for topical insulin 1 unit/drop four times daily and standard artificial tears in treating diabetics with dry eye disease”.

### Quality and Bias of Included Studies

#### Quality Distribution

Among the ten included articles, three exhibited robust methodology, warranting categorization as “good quality” Three others fell under the “moderate quality” category, while the remaining four displayed methodological limitations, leading to a “low quality” classification.

#### Risk of Bias Assessment

The QUADAS-2 tool revealed a concerning distribution of bias risk. Four studies demonstrated a high likelihood of bias, potentially influencing their findings. Three studies exhibited moderate bias risk, requiring cautious interpretation. Fortunately, three studies displayed a low risk of bias, offering a stronger foundation for their conclusions.

The synthesis of the encompassed research and corresponding results is depicted in Table 1.

**Table 1.**
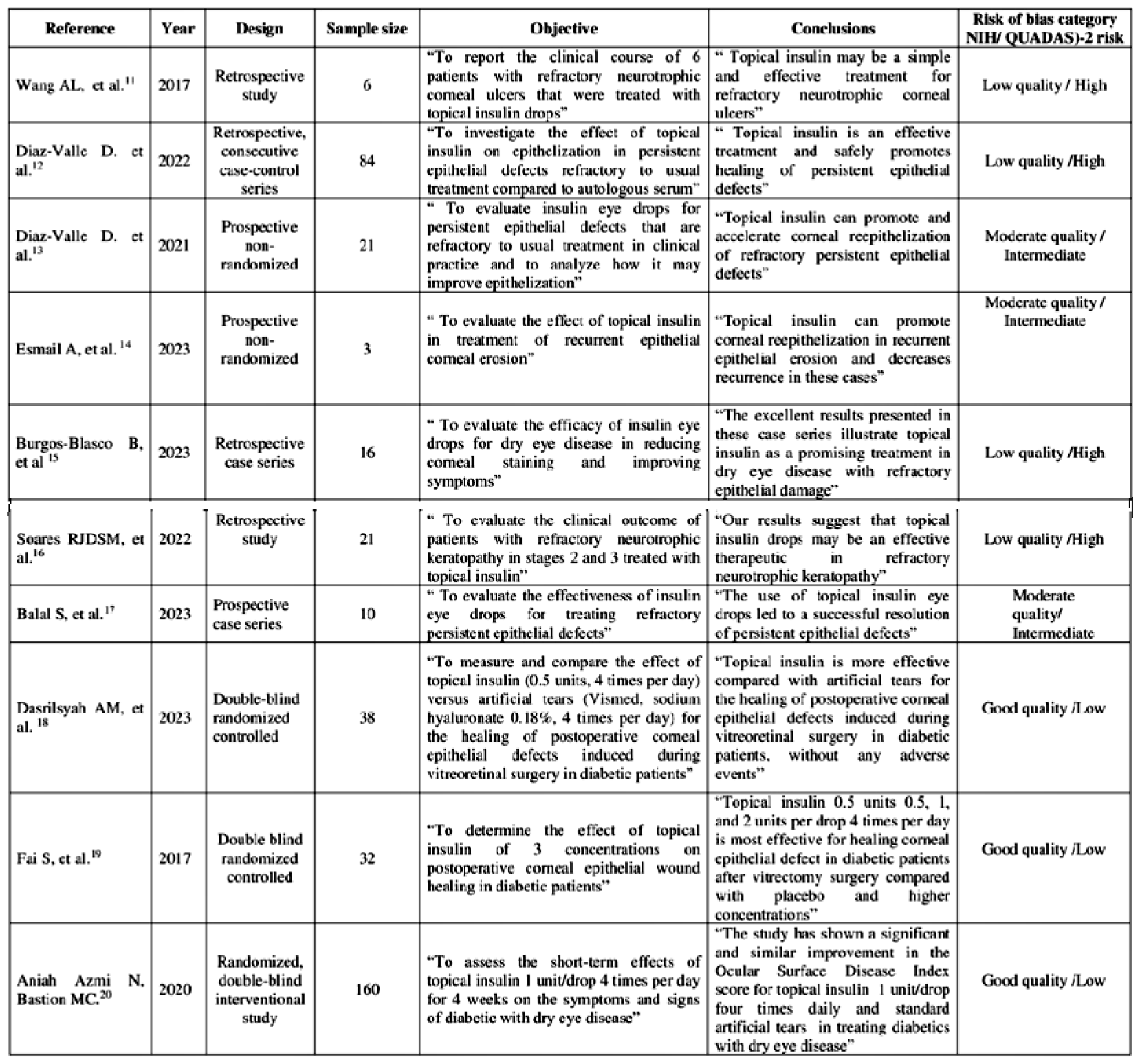
The synthesis of the research and corresponding results.

### Safety and Efficacy of Topical Insulin for Ocular Disease

The ten studies evaluated in this systematic review highlighted the potential of ocular topical insulin as a simple, safe, and cost-effective treatment for various ocular conditions, such as neurotrophic corneal ulcers, corneal reepithelization of persistent epithelial defects, dry eye disease, recurrent epithelial erosion, and corneal epithelial defects induced during vitreoretinal surgery in diabetic patients, without any adverse events. These results suggest that topical ocular insulin could be a promising first-line treatment for a number of ocular conditions and is well tolerated, readily available, and economical, potentially offering advantages over conventional treatments.

## DISCUSSION

Despite its established role in systemic glycemic management, the application of insulin topically to the eye for the treatment of ocular diseases remains a relatively novel investigative avenue. The ten studies evaluated in this systematic review highlighted the potential of ocular topical insulin as a simple, safe, and cost-effective treatment for various ocular conditions. Thus, by critically analyzing the available data, we emphasize the potential of this promising therapeutic approach and provide insights into its clinical utility.

Emerging data implicates topical insulin as a potent modulator of the wound healing cascade. Historically, insulin administration has been primarily utilized for the treatment and management of diabetes mellitus. However, recent research has unveiled the potential therapeutic applications of insulin in various other medical fields, warranting further investigation. The insulin pleiotropic effects extend beyond glycemic control, exhibiting growth factor-like activity.^21^ This translates to enhanced keratinocyte migration, fibroblast proliferation, and extracellular matrix protein deposition, ultimately promoting tissue regeneration. Furthermore, topical insulin modulates the inflammatory milieu, influencing the release of pro- and anti-inflammatory cytokines to orchestrate a balanced healing response. While the exact mechanisms remain under active investigation, the potential of topical insulin as a therapeutic adjunct in wound management warrants further exploration.^22^

Christie and Hanzal’s pioneering work in 1931 explored topical ocular insulin administration for controlling serum glucose levels.^23^ However, subsequent studies have consistently demonstrated the ineffectiveness of such formulations (insulin in isotonic sodium chloride solutions) in modulating systemic D-glucose levels in both humans and animals.^24^ The current findings concur with this established knowledge, revealing no significant impact of topical insulin on serum glucose levels. This lack of systemic effect suggests that the observed improvement in diabetic corneal re-epithelialization rates likely occurs at the cellular level, rather than through any broader systemic influence. This is further supported by the presence of glucose transporter 1 (GLUT 1) in the corneal epithelium, which facilitates glucose uptake independently of insulin.^25^ While the precise mechanism of topical insulin in ocular disease remains elusive, compelling evidence points towards its potential involvement in corneal nerve regeneration and enhanced epithelial cell migration.^26^ In vitro models of corneal epithelial, wound healing further support this notion, demonstrating the ability of topical insulin to stimulate cell migration and facilitate artificial wound closure in cultured corneal epithelial sheets.^22^

Systematic literature reviews play a critical role in advancing medical knowledge by providing a comprehensive and unbiased synthesis of available research.^27^ In the field of ophthalmology, where there is a constant influx of new information, systematic literature reviews are particularly valuable for informing clinical practice and guiding future research.^28^ Our systematic review on the safety and efficacy of topical insulin for ocular disease addresses a significant knowledge gap in this area. To date, no other systematic literature reviews has been published on this topic, which highlights the importance of our work.

The quality of articles included in a systematic review is paramount for determining the reliability of its findings. Assessing the risk of bias is a crucial component in ensuring the review’s quality. The presence of bias and varying quality levels in the included studies necessitates careful consideration when interpreting the overall findings. Studies classified as “good quality” and “low risk of bias” provides more reliable evidence, while those with lower quality and higher bias risk necessitate cautious interpretation and potentially require corroboration from additional high-quality studies. In this review, we utilized two validated tools to assess the risk of bias: the NIH tool and the QUADAS-2.^29,10^ The NIH tool assesses the risk of bias in observational studies, while the QUADAS-2 evaluates the risk of bias in intervention studies. Evaluating the risk of bias allowed us to identify studies with a high risk of bias and, consequently, lower reliability. This assessment contributed to the selection of high-quality studies for data synthesis.

Our systematic review examined ten studies exploring the efficacy of topical insulin for various ocular conditions. The studies employed diverse methodologies, ranging from retrospective case series to double-blind randomized controlled trials. Despite methodological differences, a consistent theme emerged: topical insulin exhibited promising outcomes across various indications. Specifically, studies demonstrated the potential of topical insulin in: Promoting healing of refractory neurotrophic corneal ulcers; Accelerating reepithelialization in persistent epithelial defects; Reducing recurrence of recurrent epithelial erosions; Improving symptoms and reducing corneal staining in dry eye disease; Enhancing postoperative corneal epithelial wound healing in diabetic patients. These findings suggest that topical insulin may be a valuable tool in managing diverse ocular pathologies, particularly in cases refractory to conventional therapies.

## CONCLUSION

This systematic review provides encouraging evidence for the therapeutic potential of topical insulin in various ocular conditions. While further research is warranted to confirm these findings in larger, well-designed studies, the current body of literature suggests that topical insulin may offer a safe and effective treatment option for patients with refractory corneal disorders. Future research should explore optimal dosing regimens, long-term safety profiles, and broader applications in ophthalmology.

## Data Availability

All data produced in the present work are contained in the manuscript

